# Religiosity and stigmatization related to mental illness among African-Americans and Black immigrants: cross-sectional observational study and moderation analysis

**DOI:** 10.1101/2021.11.02.21265783

**Authors:** Aderonke Bamgbose Pederson, Valerie A. Earnshaw, Roberto Lewis-Fernández, Devan Hawkins, Dorothy I. Mangale, Alexander C. Tsai, Graham Thornicroft

**Affiliations:** Department of Psychiatry, Massachusetts General Hospital, Harvard Medical School, Boston, Massachusetts; Department of Human Development and Family Sciences, University of Delaware, Newark, Delaware; Columbia University College of Physicians and Surgeons and New York State Psychiatric Institute, New York, New York; Public Health Program, School of Arts and Sciences, Massachusetts College of Pharmacy and Health Sciences, Boston, Massachusetts; Department of Global Health, University of Washington, Seattle, Washington; Center for Global Health and Mongan Institute, Massachusetts General Hospital, Harvard Medical School, Boston, Massachusetts; Harvard Medical School, Boston, Massachusetts; Centre for Global Mental Health and Centre for Implementation Science, Institute of Psychiatry, Psychology and Neuroscience, King’s College London, United Kingdom

**Keywords:** Mental health, religiosity, stigma, social contact, Black adults

## Abstract

**Objectives:** Stigma about mental illness is a known barrier to engagement in mental health services. This study aimed to estimate the associations between religiosity and mental illness stigma among Black adults.

**Design:** We conducted an online cross-sectional study of Black adults in the United States (n=269, ages 18-65) from diverse ethnic backgrounds.

**Results:** Most (n=248 [92%]) participants attended religious services; while 21 (8%) never attended. Social distance was assessed as an index of past or current stigmatizing behavior. After adjusting for demographic factors, respondents with higher attendance at religious services or greater engagement in religious activities (e.g., prayer, meditation or Bible study) reported greater proximity to people living with mental health problems (RR=1.72; 95% CI: 1.14, 2.59 and RR=1.82; CI: 1.18, 2.79, respectively). Despite reporting greater past or current social proximity, respondents with higher religiosity indices also reported greater future intended stigmatizing behavior (or lower future intended social proximity) (RR’s=0.92-0.98). Ethnicity moderated the association between religiosity and future intended stigmatizing behavior. Black immigrants with higher religiosity reported lower future intended stigmatizing behavior (RR=1.16 CI: 1.02 – 1.32) whereas African-Americans with higher religiosity reported greater future intended stigmatizing behavior (RR = 0.83 CI: 0.76, 0.91).

**Conclusions:** Higher indices of religiosity were associated with lower past or current stigmatizing behavior towards individuals living with mental health problems but not lower future intended stigmatizing behavior. Focusing specifically on future intended stigmatizing behavior and the respondent’s level of religiosity, age, and ethnicity may be critical for designing effective stigma-reducing interventions for Black adults.

## Introduction

Black adults comprise 12% of the general US population and experience a disproportionately high burden of mental illness (e.g. more chronic illness) compared to white adults (Breslau et al., 2006; Ward et al., 2013). The Black population has grown by over 10 million people since 2000, a 29% increase over two decades (Tamir, 2021). The Black immigrant population -- 88% of whom come from Africa or the Caribbean (Tamir, 2021) -- has grown even more rapidly. Black immigrants are often overlooked in health research because studies usually ignore the diversity in nativity and ethnic origin within the Black population (Mehta et al., 2015; Pinder et al., 2016). Identifying barriers to engagement in and adherence to mental health treatment among Black adults, including Black immigrants, is critical for improving their mental health care and eliminating inequities in mental health in the US.

Past research suggests that stigma negatively affects engagement in mental health services, including treatment adherence (Corrigan, 2004; Corrigan et al., 2014; Rao et al., 2007; Vinson et al., 2016; Ward et al., 2013). Stigma has been conceptualized as a broad social process that emerges at the co-occurrence of labeling, stereotyping, separation, status loss, and discrimination, and is based on a power structure in which the stigmatized group has less power (Link & Phelan, 2006; Lucas & Phelan, 2012). *Mental illness stigma* refers to negative knowledge, attitudes, beliefs, or behaviors towards people with mental illness and mental health services (Corrigan et al., 2012). The desire to distance oneself socially from individuals with mental illness is a manifestation of negative attitudes towards them and can lead to stigma-related discriminatory behaviors (Corrigan et al., 2012; Evans-Lacko et al., 2011; Wang et al., 2019).

The relationship between stigma and mental illness in the Black community is complex. Black individuals with mental illness may be stigmatized because of multiple intersecting identities, including their racial identity, mental health problems, and migrant status in the case of Black immigrants (Stangl et al., 2019). They may also perpetuate stigma towards mental illness; for example a national survey showed 63% of Black people believe depression is a personal weakness (Ward et al., 2013). Their combined experiences as potentially both stigmatized and stigmatizer contributes to low utilization of mental health services and therefore to poor mental health outcomes among Black individuals. Compared with white adults, Black adults endorse more stigma towards mental illness and have more chronic and persistent mental illness, including higher levels of associated disability (Snowden, 2001; Ward et al., 2009; Ward et al., 2013; Williams et al., 2007). This complexity shows that the balance between holding stigmatizing views about mental illness and experiencing the stigma of mental illness is likely to be influenced by one’s cultural context (Lawrence Hsin Yang et al., 2014).

Religious and cultural beliefs can influence conceptualizations of mental health, help-seeking, and treatment outcomes, especially in the Black community, where religious organizations serve as a focal point of support (Nguyen, 2020). Black people with higher religiosity are less likely to seek mental health services and more likely to prefer mental health support from religious bodies (Lukachko et al., 2015; Maura & Weisman de Mamani, 2017). Mental illness stigma may help explain this finding. To address the impact of mental illness stigma on the Black community and reduce barriers to care, mental health systems would benefit from clarifying the relationships between religiosity and stigma in the Black community while taking into account its internal diversity (Lacey et al., 2016). Understanding this association will facilitate the design of targeted, effective interventions to reduce mental illness stigma and its adverse consequences such as delayed treatment seeking (John R. Peteet, 2019). However, the association between religiosity and stigma remains understudied in the Black community (John R. Peteet, 2019; L. H. Yang et al., 2014).

Few studies address the relationship between religiosity and use of mental health services in underserved ethnoracial communities (Lukachko et al., 2015). Most studies on religion and mental health focus on how religiosity provides social support and confers resilience (Li et al., 2016; VanderWeele et al., 2016), especially among disadvantaged or underserved communities (Nguyen, 2020). Our study adds to this growing body of knowledge by focusing on religiosity and mental illness stigma, both of which may influence mental health service use. We examined the association between religiosity and mental health-related behavioral discrimination (Evans-Lacko et al., 2011). Our primary outcomes assess reported (past or current) discriminatory behavior towards individuals with mental health problems as well as intended (future) discriminatory behavior. Most studies on mental illness stigma-reducing interventions assess knowledge and attitudes primarily, although research has emphasized that the elimination of discriminatory behavior towards mental illness is the most important factor in combating mental illness stigma (Corrigan, 2012; Graham Thornicroft et al., 2016). Moreover, the primary method to reduce stigma is through contact-based interventions, which are based on the theory that positive and voluntary interaction with people who have mental health problems reduces discriminatory behavior and other enactments of stigma (Corrigan et al., 2012; Mousa, 2020; Paluck et al., 2016; Thornicroft, 2006; Graham Thornicroft et al., 2016). However, previous studies usually do not address the time course of contact behavior, such as how past/current behavior may differ from future intended behavior based on the quality of past experiences. The aim of this study is to examine the associations between religiosity and stigmatizing behavior in two time frames (past/current vs. future) among Black adults in the U.S., and to determine the extent to which these associations differed by ethnicity (African-Americans, African immigrants, and Afro-Caribbean immigrants). Insights gained from examining religiosity and its association with mental illness stigmatizing behavior in the Black community can enhance the effectiveness of stigma-reduction interventions designed to optimize utilization of mental health services.

## Methods

### Study design, setting, and participants

An online cross-sectional survey was conducted in September-October 2020. African-Americans, African immigrants, and Afro-Caribbean immigrants (n=269) in the United States were recruited using social media (email, Facebook, Twitter) in partnership with local community-based organizations (World Relief Chicago, the United African Organization, Refugee One), which shared the survey with their members. The eligibility criteria included: identifying as Black, African-American, African or Afro-Caribbean; ages 18-65; residing in the US; and English-language fluency. Additional demographic questions were asked about participant ethnicity and race.

### Procedures

All data were elicited using an online survey.

### Ethical Review

Study procedures were approved by the Institutional Review Board at XXXX. All participants provided written informed consent.

### Measures

Study participants were asked to report their age, gender, race, ethnicity, marital status, education, annual income, insurance status, citizenship status, and length of stay in the US.

#### Primary exposure

##### religiosity

To assess religiosity, we used the Duke University Religion Index (DUREL), a five-item measure that assesses frequency of attending religious services (1 item); frequency of private religious activities, e.g. prayer, meditation, or Bible study (1 item); and intrinsic religiosity, based on one’s experience of the presence of the divine, whether religiosity influences one’s approach to life, and whether one’s religion is carried into other life activities (3 items) (Koenig & Büssing Arndt, 2010). Responses are based on a 5-point Likert or 6-point Likert-type scale. Responses for subscales 1 (attending religious services) and 2 (engaging in private religious activities) range from *never* to *more than once a week* for subscale 1 and *more than once a day* for subscale 2. Responses for subscale 3 (*intrinsic religiosity*) range from *definitely not true* to *definitely true*. The DUREL scale has high test-retest and internal consistency reliability (Cronbach’s alpha = 0.78-0.91) (Koenig & Büssing Arndt, 2010). The scale is highly convergent with other measures of religiosity such as Hoge’s Intrinsic Religiosity Scale and the Allport and Ross Intrinsic-Extrinsic scale (r’s = 0.71-0.86) (Koenig & Büssing Arndt, 2010).

#### Primary outcome

##### mental illness stigma (social distancing)

To assess mental illness stigma, we used the Reported and Intended Behavior Scale (RIBS). The RIBS is an 8-item scale that measures reported (past and current) behavior and intended (future) behavior regarding people with mental health problems (Evans-Lacko et al., 2011). It is composed of two groups of items. The first four items assess past and current social distance: “are you currently or have you ever…” “lived,” “worked,” “been a neighbor to” or “had a close friendship with…” “someone with a mental health problem.” Responses include *yes, no*, or *don’t know*. Based on scoring guidelines, these first four items are assessed as individual outcomes and not summed. In our sample, the Cronbach’s alpha for the first four items ranged from 0.72-0.81. The second four items assess intended future behavior as a single continuous variable, based on a summed score; high scores represent more favorable behavior (i.e., less intended stigmatizing behavior). Responses to the intended behavior measure include 5 options measured on a Likert-type scale (*agree strongly –* coded as 5 – to *disagree strongly –* coded as 1 and the option *don’t know –* coded neutrally as 3). The items are worded similarly to the first four items (e.g. “In the future I would be willing to live with someone with a mental health problem”). In our sample, the Cronbach’s alpha for the second four items, a continuous subscale, was 0.85 (10). Overall test-retest reliability is 0.75 (Evans-Lacko et al., 2011).

### Data analysis

Socio-demographic characteristics were described using summary statistics. To examine the associations between religiosity and stigmatizing behavior, we used generalized linear models. We assessed attendance at religious services and engagement in private religious activities as binary variables, comparing respondents who reported *a few times a month* or more to those who reported *a few times a year* or less. Given the sample size, we dichotomized responses to the DUREL religiosity first two subscales into (1) those who responded *a few times a year* or less vs. (2) those who responded *a few times a month* or more frequently rather than assessing each of the responses individually. For the relationship between the binary DUREL subscales 1 and 2 and each of the first four items of the RIBS scale, in separate models we used Poisson regression with a log link, given the binary nature of the outcome variable. From these models we derived rate ratios (RR) to assess the ratio of the probability of responding *yes* to having past or current contact with someone with mental health problems among those who reported *a few times a month* or more frequently compared to those who reported *a few times a year* or less frequently. For DUREL subscale 3, since the predictor was treated as a continuous variable, the RR represents the average change in the probability of responding *yes* per unit change in the subscale. For the relationship between the DUREL subscales and the summed RIBS subscore, gamma regression using generalized linear models was used, given the continuous outcome measure. Similar definitions for the RR were used as in the analyses above.

To account for confounding, we adjusted for age (under-35 vs. 35+), education (high school education or less vs. some college or more), and ethnicity (Black Immigrant vs. African-American). Afro-Caribbean immigrants (n=13) were combined with African immigrants (n=52) into a *Black immigrant* group due to their small sample size. There is precedent for this, given similarities in migration experience among individuals who identify as Black (Amuta-Jimenez et al., 2019). To check for effect modification, the same regressions described above were performed with stratification by age, education, and ethnicity. For all models, statistical significance was assessed as p≤0.05; SAS Version 9.3 (SAS Institute Inc., Cary, NC) was used throughout.

## Results

Of 269 participants (Table 1), 59.85% were 34 or younger, 57.25% identified as male, and 63.94% were married. In addition, 32.35% had no college education, 43.13% reported a personal income less than $30,000, and 55.39% had public insurance. Participants identified as African-American (75.46%), African (19.33%), and Afro-Caribbean (4.83%). Over half (56.88%) had lived in the United States for more than 10 years. Three-quarters (76.21%) were US citizens.

**Table 1.** Socio-demographic characteristics of the sample (N=269)

|  | n | % |
| --- | --- | --- |
| Age |  |  |
| 18-34 | 161 | 59.85 |
| 35-65 | 108 | 40.15 |
| Gender |  |  |
| Male | 154 | 57.25 |
| Female | 115 | 42.75 |
| Marital status |  |  |
| Married | 172 | 63.94 |
| Unmarried, living with a romantic partner | 45 | 16.73 |
| Never married | 29 | 10.78 |
| Separated | 7 | 2.60 |
| Divorced | 15 | 5.58 |
| Widowed | 1 | 0.37 |
| Education |  |  |
| Less than high school | 5 | 1.86 |
| High school diploma / GED | 44 | 16.36 |
| Trade school/vocational school | 38 | 14.13 |
| Some college, no degree | 91 | 33.83 |
| 2-year college | 44 | 16.36 |
| 4-year college degree | 40 | 14.87 |
| Master's degree | 4 | 1.49 |
| Doctoral degree | 3 | 1.12 |
| Annual income |  |  |
| \$9,999 or less | 15 | 5.58 |
| \$10,000 to \$29,999 | 101 | 37.55 |
| \$30,000 to \$49,999 | 119 | 44.24 |
| \$50,000 or more | 34 | 12.64 |
| Insurance |  |  |
| None | 60 | 22.30 |
| Public | 149 | 55.39 |
| Private | 60 | 22.30 |
| Ethnicity |  |  |
| African | 52 | 19.33 |
| African-American | 203 | 75.46 |
| Afro-Caribbean | 13 | 4.83 |
| Other* | 1 | 0.37 |
| How long in U.S.? |  |  |
| Less than 2 years | 6 | 2.23 |
| 2 to 5 years | 37 | 13.75 |
| 6 to 10 years | 73 | 27.14 |
| More than 10 years | 154 | 56.88 |
| Citizenship status |  |  |
| Citizen | 205 | 76.21 |
| Non-citizen | 64 | 23.79 |

\*Person identified as Black but did not select any of the three ethnicities and was not included in the stratified analysis that measured ethnicity

As shown in Table 2, the sample was highly religious, 75.5% reported having attended religious services a few times per year or more and 53.2% reported having engaged in religious activities at least once per week or more.

**Table 2.**
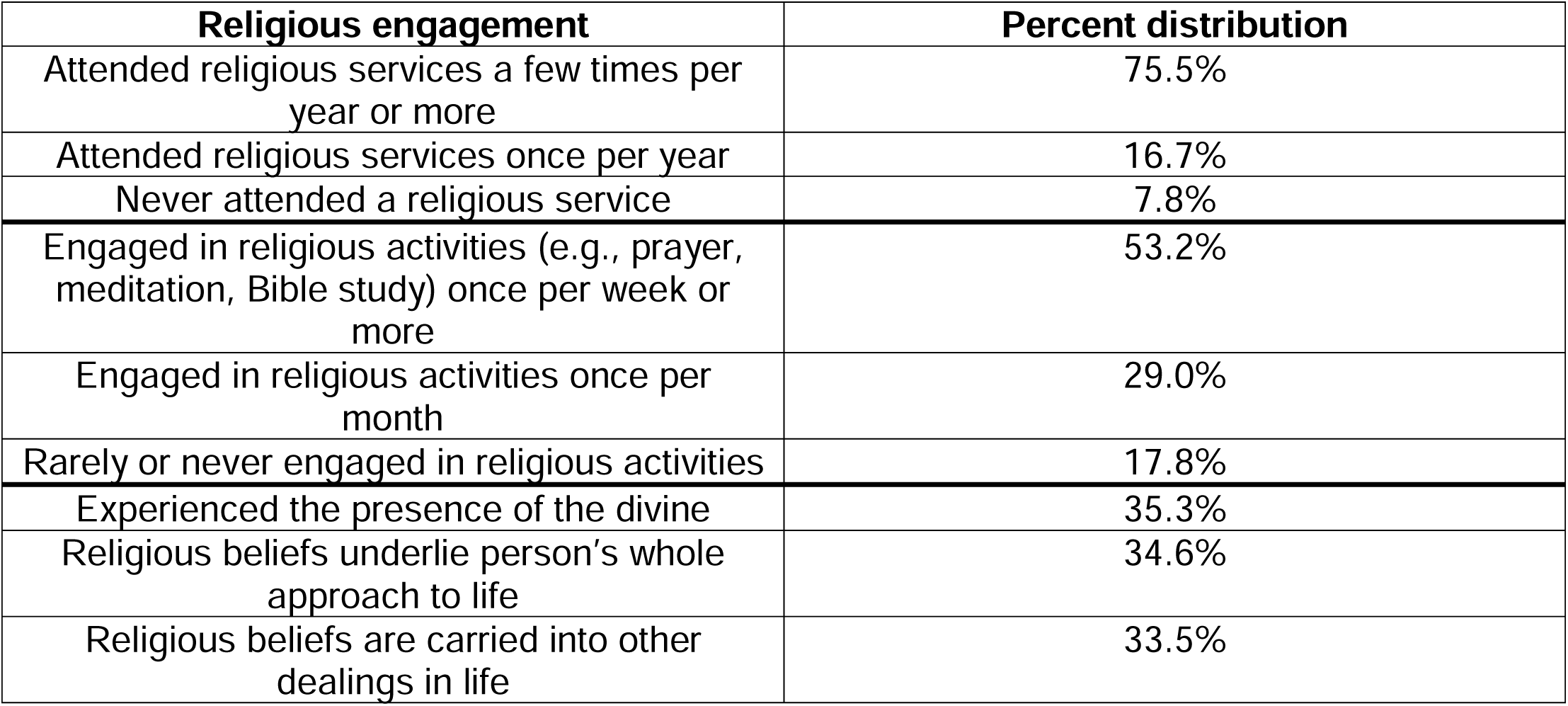
Religiosity and level of engagement in religious activities (N=269)

| **Religious engagement** | **Percent distribution** |
| --- | --- |
| Attended religious services a few times per year or more | 75.5% |
| Attended religious services once per year | 16.7% |
| Never attended a religious service | 7.8% |
| Engaged in religious activities (e.g., prayer, meditation, Bible study) once per week or more | 53.2% |
| Engaged in religious activities once per month | 29.0% |
| Rarely or never engaged in religious activities | 17.8% |
| Experienced the presence of the divine | 35.3% |
| Religious beliefs underlie person's whole approach to life | 34.6% |
| Religious beliefs are carried into other dealings in life | 33.5% |

### The association between religiosity and reported stigma towards persons with mental health problem

Adjusting for age, education, and ethnicity, we examined the association between religiosity and reported social distance from persons with mental health problems as well as future intended stigmatizing behavior. Table 3 shows the results of the unadjusted (Model 1) and adjusted (Model 2) analyses. Statistically significant findings were generally unchanged between the two models.

**Table 3.** Unadjusted (Model 1) and adjusted (Model 2) analyses examining the association between indices of religiosity (Duke University Religion Index) and reported past and current stigmatizing behavior as well as future intended stigmatizing behavior (Reported and Intended Behavior Scale) (N=269)

|  | Social Distance (Reported and Intended) |  |  |  |  |  |  |  |  |  |  |  |  |  |  |  |  |  |  |  |
| --- | --- | --- | --- | --- | --- | --- | --- | --- | --- | --- | --- | --- | --- | --- | --- | --- | --- | --- | --- | --- |
|  | Living with someone with mental health problems |  |  |  | Working with someone with mental health problems |  |  |  | Neighbor to someone with mental health problems |  |  |  | Close friend with someone with mental health problems |  |  |  | Future intended social distance (RIBS average) |  |  |  |
|  | Unadjusted |  | Adjusted |  | Unadjusted |  | Adjusted |  | Unadjusted |  | Adjusted |  | Unadjusted |  | Adjusted |  | Unadjusted |  | Adjusted |  |
|  | RR (95% CI) | P-value | RR (95% CI) | P-value | RR (95% CI) | P-value | RR (95% CI) | P-value | RR (95% CI) | P-value | RR (95% CI) | P-value | RR (95% CI) | P-value | RR (95% CI) | P-value | RR (95% CI) | P-value | RR (95% CI) | P-value |
| **Attendance at religious services** |  |  |  |  |  |  |  |  |  |  |  |  |  |  |  |  |  |  |  |  |
| A few times a year or less (ref) | 1 |  | 1 |  | 1 |  | 1 |  | 1 |  | 1 |  | 1 |  | 1 |  | 1 |  | 1 |  |
| A few times a month or more | **1.69**<br>(1.13, 2.55) | **0.01\*** | **1.72**<br>(1.14, 2.59) | **0.01\*** | 1.32<br>(0.89, 1.95) | 0.01 | 1.29<br>(0.87, 1.91) | 0.21 | 1.17<br>(0.77, 1.78) | 0.48 | 1.19<br>(0.78, 1.82) | 0.41 | **1.69**<br>(1.09, 2.61) | **0.02\*** | **1.66**<br>(1.07, 2.57) | **0.02\*** | 0.96<br>(0.89, 1.03) | 0.27 | 0.96<br>(0.90, 1.03) | 0.290 |
| **Private religious activities** |  |  |  |  |  |  |  |  |  |  |  |  |  |  |  |  |  |  |  |  |
| Once a week or less (ref) | 1 |  | 1 |  | 1 |  | 1 |  | 1 |  | 1 |  | 1 |  | 1 |  | 1 |  | 1 |  |
| A few times a week or more | **1.78**<br>(1.17, 2.71) | **0.007\*** | **1.82**<br>(1.18, 2.79) | **0.007\*** | 1.18<br>(0.77, 1.81) | 0.44 | 1.22<br>(0.79, 1.89) | 0.37 | 1.32<br>(0.85, 2.04) | 0.21 | 1.22<br>(0.78, 1.91) | 0.37 | 1.45<br>(0.92, 2.30) | 0.11 | 1.45<br>(0.90, 2.32) | 0.12 | **0.91**<br>(0.84, 0.98) | **0.01\*** | **0.92**<br>(0.85, 0.99) | **0.035\*** |
| **Intrinsic religiosity** | **1.16**<br>(1.08, 1.25) | **<0.0001\*** | **1.16**<br>(1.08, 1.25) | **<0.0001\*** | 1.04<br>(0.98, 1.11) | 0.19 | 1.04<br>(0.97, 1.11) | 0.23 | 1.01<br>(0.94, 1.08) | 0.76 | 1.02<br>(0.95, 1.09) | 0.67 | **1.13**<br>(1.05, 1.22) | **0.002\*** | **1.13**<br>(1.05, 1.22) | **0.002\*** | **0.98**<br>(0.97, 0.99) | **0.002\*** | **0.98**<br>(0.97, 0.99) | **0.002\*** |

Poisson regression with a log link was used to calculate rate ratios (RR) to assess changes in social distance and stigma per unit change in religiosity score for each subscale (religious service attendance, private religious activities, intrinsic religiosity). For future intended stigmatizing behavior, we used gamma regression with generalized linear models. These models represent the average change in RIBS score per unit change in DUREL. Model 1 – Only DUREL scale; Model 2 – DUREL adjusting for age, education, and ethnicity; \***p < 0.05**

#### 1.1 Attendance at religious meetings (church, synagogue, or other) was associated with social distance from persons with mental health problems

As in the unadjusted analysis, after adjusting for age, education, and ethnicity, respondents who attended religious services more frequently were 72% more likely to report ever living with someone with mental health problems (RR=1.72, p=0.001; CI: 1.14, 2.59) and 66% more likely to ever have a close friend with mental health problems (RR=1.66, p=0.024; CI: 1.07, 2.57). However, they were not significantly more likely to endorse future intended stigmatizing behavior.

#### 1.2 Engagement in religious activities (e.g., prayer, meditation, Bible study) was associated with social distance from persons with mental health problems

Based on the adjusted analysis, respondents who engaged in religious activities at least a few times a month were 82% more likely to report ever living with someone with mental health problems compared to respondents who engaged in religious activities less frequently (RR=1.82, p=0.007; CI: 1.18, 2.79). Respondents who engaged in religious activities more frequently also had a mean RIBS score that was 8% lower (RR=0.92, p=0.035; CI: 0.85, 0.99), indicating greater future intended stigmatizing behavior.

#### 1.3 Belief that religion has a central role (intrinsic religiosity) and its association with past, current or future social distance and behavior related to persons with mental health problems

Based on adjusted analyses, respondents with greater intrinsic religiosity were 16% more likely to report ever living with someone with mental health problems (RR=1.16, p<0.0001; CI: 1.08, 1.25) and 13% more likely to ever having a close friend with mental health problems (RR=1.13, p=0.002; CI: 1.05, 1.22). However, they had an average stigma score only 2% lower than respondents with low intrinsic religiosity (RR=0.98, p=0.002; CI: 0.97, 0.99), indicating slightly higher future intended stigmatizing behavior.

**Table 4a.** Association between indices of religiosity (Duke University Religion Index) and future intended stigma behavior (Reported and Intended Behavior Scale) stratified by effect modifiers (ethnicity, age, education) to assess moderation.

|  | Future intended social distance (RIBS average) |  |  |  |
| --- | --- | --- | --- | --- |
|  | 18-34 |  | 35-65 |  |
| Attendance at religious services | RR (95% CI) | P-value | RR (95% CI) | P-value |
| A few times a year or less | 1 |  | 1 |  |
| A few times a month or more | 0.99 (0.89, 1.09) | 0.8 | 0.92 (0.84, 1.02) | 0.1111 |
| Private religious activities |  |  |  |  |
| Once a week or less | 1 |  | 1 |  |
| A few times a week or more | **0.90 (0.81, 0.99)** | **0.026\*** | 0.96 (0.84, 1.09) | 0.530 |
| Intrinsic religiosity | 0.98 (0.96, 1.00) | 0.018 | 0.98 (0.97, 1.00) | 0.020 |
|  | High school or less |  | Some college or more |  |
| Attendance at religious services | RR (95% CI) | P-value | RR (95% CI) | P-value |
| A few times a year or less | 1 |  | 1 |  |
| A few times a month or more | 0.94 (0.80, 1.09) | 0.406 | 0.97 (0.90, 1.04) | 0.410 |
| Private religious activities |  |  |  |  |
| Once a week or less | 1 |  | 1 |  |
| A few times a week or more | **0.78 (0.67, 0.91)** | **0.001\*** | 0.99 (0.91, 1.08) | 0.821 |
| Intrinsic religiosity | **0.97 (0.95, 0.99)** | **0.003\*** | 0.99 (0.98, 1.00) | 0.109 |
|  | Black immigrant |  | African-American |  |
| Attendance at religious services | RR (95% CI) | P-value | RR (95% CI) | P-value |
| A few times a year or less | 1 |  | 1 |  |
| A few times a month or more | 1.11 (0.98, 1.26) | 0.103 | 0.92 (0.84, 1.00) | 0.04 |
| Private religious activities |  |  |  |  |
| Once a week or less | 1 |  | 1 |  |
| A few times a week or more | **1.16 (1.02, 1.32)** | **0.022\*** | **0.83 (0.76, 0.91)** | **<0.0001\*** |
| Intrinsic religiosity | 1.01 (0.99, 1.04) | 0.215 | **0.97 (0.96, 0.99)** | **<0.0001\*** |

**Table 4b.**
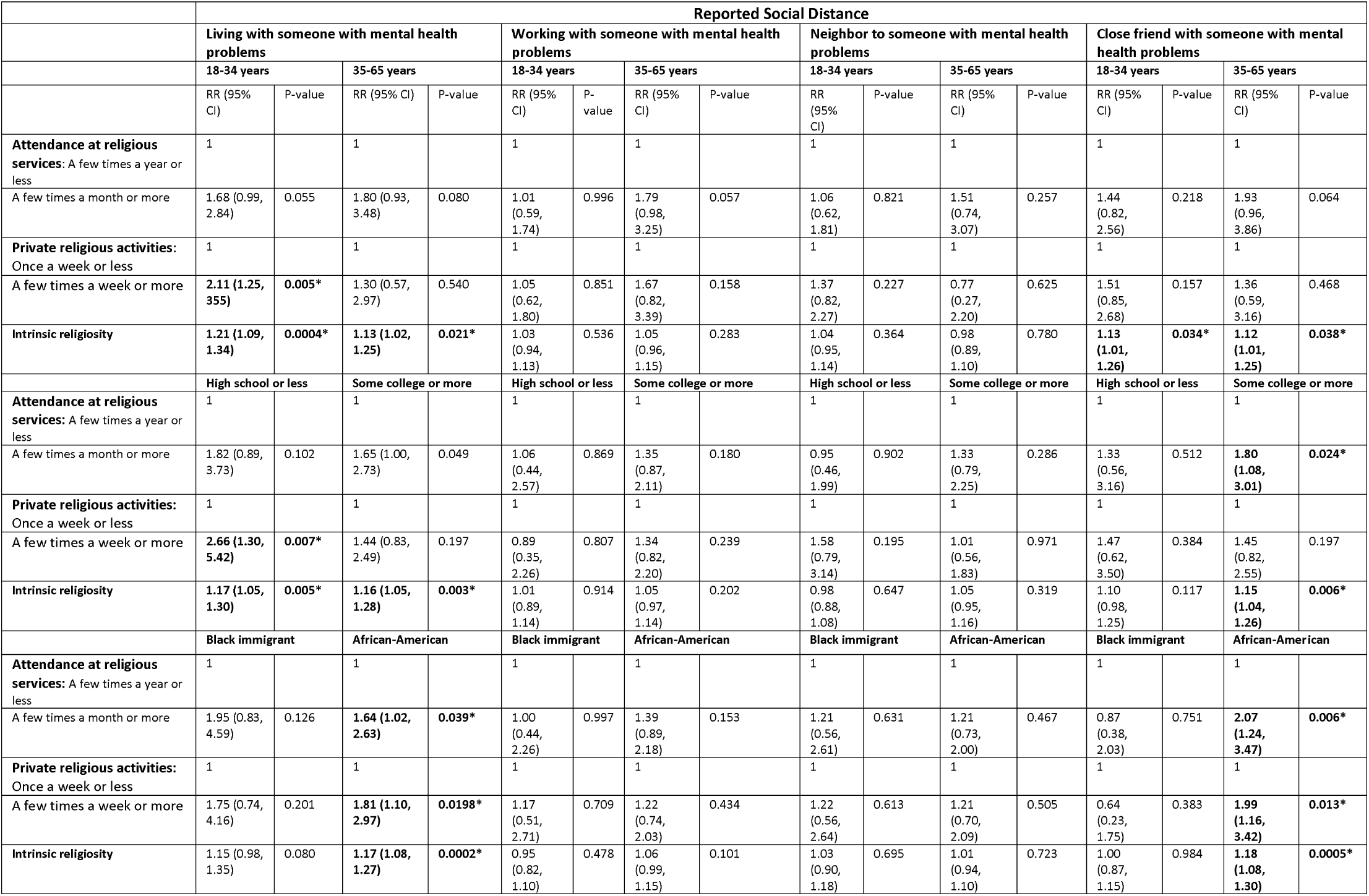
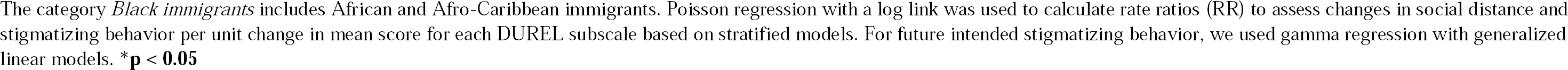
Association between indices of religiosity (Duke University Religion Index) and reported current or past stigmatizing behavior (Reported and Intended Behavior Scale) stratified by effect modifiers (ethnicity, age, education) to assess moderation.

|  | Reported Social Distance |  |  |  |  |  |  |  |  |  |  |  |  |  |  |  |
| --- | --- | --- | --- | --- | --- | --- | --- | --- | --- | --- | --- | --- | --- | --- | --- | --- |
|  | Living with someone with mental health problems |  |  |  | Working with someone with mental health problems |  |  |  | Neighbor to someone with mental health problems |  |  |  | Close friend with someone with mental health problems |  |  |  |
|  | 18-34 years |  | 35-65 years |  | 18-34 years |  | 35-65 years |  | 18-34 years |  | 35-65 years |  | 18-34 years |  | 35-65 years |  |
|  | RR (95% CI) | P-value | RR (95% CI) | P-value | RR (95% CI) | P-value | RR (95% CI) | P-value | RR (95% CI) | P-value | RR (95% CI) | P-value | RR (95% CI) | P-value | RR (95% CI) | P-value |
| **Attendance at religious services:** A few times a year or less | 1 |  | 1 |  | 1 |  | 1 |  | 1 |  | 1 |  | 1 |  | 1 |  |
| A few times a month or more | 1.68 (0.99, 2.84) | 0.055 | 1.80 (0.93, 3.48) | 0.080 | 1.01 (0.59, 1.74) | 0.996 | 1.79 (0.98, 3.25) | 0.057 | 1.06 (0.62, 1.81) | 0.821 | 1.51 (0.74, 3.07) | 0.257 | 1.44 (0.82, 2.56) | 0.218 | 1.93 (0.96, 3.86) | 0.064 |
| **Private religious activities:** Once a week or less | 1 |  | 1 |  | 1 |  | 1 |  | 1 |  | 1 |  | 1 |  | 1 |  |
| A few times a week or more | **2.11 (1.25, 3.55)** | **0.005\*** | 1.30 (0.57, 2.97) | 0.540 | 1.05 (0.62, 1.80) | 0.851 | 1.67 (0.82, 3.39) | 0.158 | 1.37 (0.82, 2.27) | 0.227 | 0.77 (0.27, 2.20) | 0.625 | 1.51 (0.85, 2.68) | 0.157 | 1.36 (0.59, 3.16) | 0.468 |
| **Intrinsic religiosity** | **1.21 (1.09, 1.34)** | **0.0004\*** | **1.13 (1.02, 1.25)** | **0.021\*** | 1.03 (0.94, 1.13) | 0.536 | 1.05 (0.96, 1.15) | 0.283 | 1.04 (0.95, 1.14) | 0.364 | 0.98 (0.89, 1.10) | 0.780 | **1.13 (1.01, 1.26)** | **0.034\*** | **1.12 (1.01, 1.25)** | **0.038\*** |
|  | High school or less |  | Some college or more |  | High school or less |  | Some college or more |  | High school or less |  | Some college or more |  | High school or less |  | Some college or more |  |
| **Attendance at religious services:** A few times a year or less | 1 |  | 1 |  | 1 |  | 1 |  | 1 |  | 1 |  | 1 |  | 1 |  |
| A few times a month or more | 1.82 (0.89, 3.73) | 0.102 | 1.65 (1.00, 2.73) | 0.049 | 1.06 (0.44, 2.57) | 0.869 | 1.35 (0.87, 2.11) | 0.180 | 0.95 (0.46, 1.99) | 0.902 | 1.33 (0.79, 2.25) | 0.286 | 1.33 (0.56, 3.16) | 0.512 | **1.80 (1.08, 3.01)** | **0.024\*** |
| **Private religious activities:** Once a week or less | 1 |  | 1 |  | 1 |  | 1 |  | 1 |  | 1 |  | 1 |  | 1 |  |
| A few times a week or more | **2.66 (1.30, 5.42)** | **0.007\*** | 1.44 (0.83, 2.49) | 0.197 | 0.89 (0.35, 2.26) | 0.807 | 1.34 (0.82, 2.20) | 0.239 | 1.58 (0.79, 3.14) | 0.195 | 1.01 (0.56, 1.83) | 0.971 | 1.47 (0.62, 3.50) | 0.384 | 1.45 (0.82, 2.55) | 0.197 |
| **Intrinsic religiosity** | **1.17 (1.05, 1.30)** | **0.005\*** | **1.16 (1.05, 1.28)** | **0.003\*** | 1.01 (0.89, 1.14) | 0.914 | 1.05 (0.97, 1.14) | 0.202 | 0.98 (0.88, 1.08) | 0.647 | 1.05 (0.95, 1.16) | 0.319 | 1.10 (0.98, 1.25) | 0.117 | **1.15 (1.04, 1.26)** | **0.006\*** |
|  | Black immigrant |  | African-American |  | Black immigrant |  | African-American |  | Black immigrant |  | African-American |  | Black immigrant |  | African-American |  |
| **Attendance at religious services:** A few times a year or less | 1 |  | 1 |  | 1 |  | 1 |  | 1 |  | 1 |  | 1 |  | 1 |  |
| A few times a month or more | 1.95 (0.83, 4.59) | 0.126 | **1.64 (1.02, 2.63)** | **0.039\*** | 1.00 (0.44, 2.26) | 0.997 | 1.39 (0.89, 2.18) | 0.153 | 1.21 (0.56, 2.61) | 0.631 | 1.21 (0.73, 2.00) | 0.467 | 0.87 (0.38, 2.03) | 0.751 | **2.07 (1.24, 3.47)** | **0.006\*** |
| **Private religious activities:** Once a week or less | 1 |  | 1 |  | 1 |  | 1 |  | 1 |  | 1 |  | 1 |  | 1 |  |
| A few times a week or more | 1.75 (0.74, 4.16) | 0.201 | **1.81 (1.10, 2.97)** | **0.0198\*** | 1.17 (0.51, 2.71) | 0.709 | 1.22 (0.74, 2.03) | 0.434 | 1.22 (0.56, 2.64) | 0.613 | 1.21 (0.70, 2.09) | 0.505 | 0.64 (0.23, 1.75) | 0.383 | **1.99 (1.16, 3.42)** | **0.013\*** |
| **Intrinsic religiosity** | 1.15 (0.98, 1.35) | 0.080 | **1.17 (1.08, 1.27)** | **0.0002\*** | 0.95 (0.82, 1.10) | 0.478 | 1.06 (0.99, 1.15) | 0.101 | 1.03 (0.90, 1.18) | 0.695 | 1.01 (0.94, 1.10) | 0.723 | 1.00 (0.87, 1.15) | 0.984 | **1.18 (1.08, 1.30)** | **0.0005\*** |

The category *Black immigrants* includes African and Afro-Caribbean immigrants. Poisson regression with a log link was used to calculate rate ratios (RR) to assess changes in social distance and stigmatizing behavior per unit change in mean score for each DUREL subscale based on stratified models. For future intended stigmatizing behavior, we used gamma regression with generalized linear models. \* $p < 0.05$

We next examined the role of age, education, and ethnicity as potential effect modifiers of the relationship between religiosity and past and current social distance as well as between religiosity and future intended stigmatizing behavior.

#### 2.1 Age (18-34 vs. 35-65)

Among participants ages 18-34, those who engaged in religious activities at least a few times a month were 111% more likely than those who engaged in religious activities less often to report ever living with someone with mental health problems, indicating lower past or current stigmatizing behavior (RR=2.11, p=0.005; CI: 1.25, 3.55). By contrast, in the same age group, those who engaged in religious activities more often had a 10% lower social distance mean score, indicating greater future intended stigmatizing behavior (RR=0.90, p=0.026; CI: 0.81, 0.99). Engagement in religious activities did not have these effects among respondents ages 35-65.

Respondents ages 18-34 with higher intrinsic religiosity reported less past or current stigmatizing behavior than individuals with lower scores: they were more likely to report ever living with someone (RR=1.21, p=0.0004; CI 1.09, 1.34) or having a close friend with mental problems (RR=1.13, p=0.034; CI: 1.01, 1.26). Similar results were observed for respondents ages 35-65 with higher intrinsic religiosity. In neither age cohort was intrinsic religiosity significantly associated with future intended stigmatizing behavior.

#### 2.2 Education (high school or less vs. some college or more)

Among respondents without any college education, those who engaged in religious activities at least a few times a month were 166% more likely than those with less-frequent religious activities to report ever living with someone with mental health problems, indicating lower past or current stigmatizing behavior (RR=2.66, p=0.007; CI: 1.30, 5.42). By contrast, in this educational category, respondents who engaged in religious activities more frequently had an average social distance score that was 22% lower than respondents with lower religious engagement, indicating greater future intended stigmatizing behavior (RR=0.78, p=0.001; CI: 0.67, 0.91).

Greater intrinsic religiosity was associated with the likelihood of ever living with someone with mental health problems equally across respondents with higher educational attainment (RR=1.16, p=0.003; CI: 1.05, 1.28) and lower educational attainment (RR=1.17, p=0.005; CI: 1.05, 1.30), indicating lower stigmatizing behavior. Individuals with higher educational attainment were also more likely to report ever having a close friend with mental health problems (RR=1.15, p=0065; CI: 1.04, 1.26). However, respondents with greater intrinsic religiosity and lower educational attainment had an average stigma score that was 3% lower than respondents with lower intrinsic religiosity, indicating slightly greater future intended stigmatizing behavior (RR=0.97, p=0.003; CI: 0.95, 0.99).

#### 2.3 Ethnicity (African-American vs. Black immigrant)

African-Americans who attended religious services frequently were more likely to report ever living with someone (RR=1.64, p=.0394; CI: 1.02, 2.63) and ever having a close friend with mental health problems (RR=2.07, p=.0056; CI: 1.24, 3.47). African-Americans who engaged in religious activities more frequently also reported similar elevations (RR=1.81, p=.0198; CI: 1.10, 2.97 and RR=1.99, p=.0128; CI: 1.16, 3.42, respectively). These significant effects were not observed among Black immigrants. However, African-Americans and Black immigrants differed in their future intended stigmatizing behavior as a function of engagement in religious activities. Among respondents with greater religious engagement, African-Americans had lower social distance scores (RR=0.83, p<0.0001; CI: 0.76, 0.91) whereas Black immigrants had higher social distance scores (RR=1.16, p=0.022; CI: 1.02, 1.32), indicating greater and lower future intended stigmatizing behavior, respectively.

Similarly, only among African-Americans, greater intrinsic religiosity was associated with ever living with someone (RR=1.17, p=.0002; CI: 1.08, 1.27) and ever having a close friend with mental health problems (RR=1.18, p=.0005; CI: 1.08, 1.30). However, among African Americans greater intrinsic religiosity was associated with a lower mean stigma score (RR=0.97, p<0.0001; CI: 0.96, 0.99), indicating slightly greater future intended stigmatizing behavior.

## Discussion

In this sample of Black respondents, we found a consistent pattern of associations between higher indices of religious involvement (higher attendance at religious services, greater engagement in religious activities, and greater intrinsic religiosity) and lower past or current social distance from individuals with mental health problems, an index of stigma behavior. Also consistently, however, participants with higher religiosity reported greater future intended stigmatizing behavior. There was one exception: among Black immigrants, greater engagement in religious activities (such as prayer or meditation) was associated with lower future intended stigmatizing behavior compared to those who were less engaged in religious activities; this pattern was not observed among African-Americans.

We hypothesize that the differential association between higher religiosity and past/current behavior vs. between higher religiosity and future intended behavior may be related to the quality of participants’ past/current contact with individuals with mental health problems. The most effective methods to reduce stigma include direct (e.g., in-person) and indirect (e.g., video-based) contact with individuals with mental illness (Patrick W. Corrigan et al., 2012; G. Thornicroft et al., 2016). However, the quality and nature of the contact are also important (Cerully et al., 2018). In stigma-reduction interventions, contact tends to occur in controlled settings and be experienced as positive and voluntary (Patrick W. Corrigan et al., 2012; G. Thornicroft et al., 2016). If contact is negative or involuntary, this may have the opposite effect on stigma, including on the willingness for future contact. It is possible that past contact with people in the community, family members, or friends that occurs in the context of an acute episode of severe mental illness or an interpersonal crisis may be perceived as negative and not associated with potential recovery (Morgan et al., 2018). This negative contact may be even more impactful when it occurs in the context of close personal relationships, such as with people one lives with and close friends. In our sample, we found an association between higher religiosity and greater proximity to people with mental health problems only in those two groups and never in the context of coworkers or neighbors. One way to understand our findings is that higher religiosity may be associated with greater proximity to individuals with mental health problems, but in contexts that intensify the negative aspects of the experience, leading to greater future intended stigmatizing behavior. Hence, the nature, quality, and setting of contact may help explain the observed discrepancy across time frames in the association between past/current and future intended stigmatizing behavior.

It is also important to understand why our data reveal a difference between Black ethnic groups in the association between higher religiosity and future intended stigmatizing behavior. Black immigrants with higher religiosity did not show greater past/current proximity to people with mental health problems and reported lower future intended stigmatizing behavior. On the other hand, African-Americans with higher religiosity showed greater past/current proximity, compared with individuals with lower religiosity, as well as more future intended stigmatizing behavior. The work of Penner and colleagues offers a possible psychological explanation based on how the development of a common, intergroup identity influences behavior and can reduce bias assessment towards others (Penner et al., 2013). The approach to intergroup needs among Black immigrants may differ from that of African-Americans. Black immigrants with higher religiosity may coalesce more strongly around their migrant status and religiosity (Hope et al., 2020), perceiving a greater affinity with other Black immigrants regardless of their mental health status. They may also interpret the mental health problems of their fellow immigrants as arising from external afflictions. These include suffering caused by supernatural powers or experiences beyond one’s control and can be associated with the expectation of healing through religious intervention, such as prayer (Conner et al., 2010; Holt et al., 2014; Holt et al., 2017; Omenka et al., 2020). In addition, Black immigrants may be less likely than African-Americans to encounter individuals with mental illness in their immigrant community due to the influence of the healthy-migrant effect (Dey & Lucas, 2006), which results in a lower likelihood of migration for individuals severely affected by mental illness. Hence, it is possible that Black immigrants with higher religiosity may be more willing to be in future contact with individuals with mental illness because of their intergroup needs, the perceived origin of the illness, the lower likelihood of negative contact with individuals with mental health problems, and the expected role of religious practices in healing illnesses.

African-Americans with higher religiosity, in contrast, may coalesce differently around their intergroup needs and willingness to engage with someone with mental health problems. Some studies show that many African-Americans may view mental illness as a weakness that is self-inflicted rather than stemming from an external source (Alang, 2016), hence dependent on will power. This more negative interpretation about past/current encounters may contribute to greater future intended stigmatizing behavior among more religious African-Americans.

Religiosity varies across age and educational level, so we adjusted for these factors in our overall analyses (Besheer Mohamed et al., 2021) and examined their impact in stratified analyses. Although greater religiosity was associated with lower past/current stigmatizing behavior in both younger and older age groups, the effect was stronger in younger adults, who reported 111% higher likelihood of ever living with someone with mental health problems currently or in the past. Younger adults with higher religious indices (and possibly higher affinity for community cohesion due to religious involvement) may experience protective effects of their religious identity on mental health. A recent meta-analysis showed that higher religiosity among youth protects against mental health problems such as substance use (Russell et al., 2020). The protective effect of religiosity on younger people may explain why younger adults reported lower past or current stigma behavior.

Participants with some college education and higher religiosity were more likely to have a friend with mental health problems. Previous studies have shown conflicting findings in the association between education and stigma: greater educational attainment was associated with both lower (Lopez et al., 2018) and greater (Alang, 2019; Lopez et al., 2018) mental illness stigma among Black adults. College students increasingly report mental health problems and there are numerous awareness initiatives focused on enhancing mental health among college students (Lattie et al., 2019). However, for both age and college education variables, lower past/current stigma was not juxtaposed with lower future stigmatizing behavior. As in the overall findings, the nature and quality of past/current contact may influence the likelihood for future contact regardless of age or educational status. Additional research is warranted to better delineate how religiosity is associated with past/current stigmatizing behavior and future stigmatizing behavior among Black people, as this may have an impact on the acceptability of stigma-reducing direct and indirect contact-based interventions.

Religious organizations constitute a trusted space in the Black community that can help promote health and wellness (Dalencour et al., 2017; Tagai et al., 2018). While some Black adults may not engage readily in mental health systems, bridging the gap between these services and Black religious communities may promote mental health care. Mental health systems should consider the role that religiosity plays in patterning social-distance behaviors between highly religious people and individuals with mental health problems, including their future intended stigmatizing behaviors and their willingness to engage in methods to reduce stigma that involve direct contact with persons with mental illness. While contact with persons with mental illness has proven effective in reducing some aspects of stigma, actual behavior change remains an ongoing challenge (G. Thornicroft et al., 2016). It is important that our stigma-reduction interventions take into account key aspects of experience that can enhance or reduce their efficacy, such as the differential impact of religiosity across ethnic groups.

### Limitations

This pilot study’s cross-sectional design prevents causal analysis of the impact of religiosity on stigma-related measures. A future study may characterize the prospective association between religiosity and social distance. Despite the moderately-large sample, which lowers the risk of random error, stratification to assess effect modification resulted in small within-group sizes even when we collapsed responses. This limited our comparisons across groups, such as comparisons across educational levels. Future studies with larger sample sizes will assess within-group differences in the association between religiosity and stigma.

## Conclusion

Our study found that higher religiosity among Black adults was associated with greater past or current social contact with individuals with mental health problems but not with greater future intended contact with them. The associations were moderated by age and ethnicity. These findings suggest that the design interventions to reduce stigma through interpersonal contact among Black adults should consider more closely the impact of religiosity, the ethnic and migration-status heterogeneity of participants, the valence (positive or negative) of past encounters with individuals with mental illness, the context (close/personal versus more formal/distant) of interactions, and the temporality (past/current versus future) of social-proximity assessments. Ongoing research is needed to further describe the mechanisms that underlie the associations between religiosity and stigma among Black adults.

## Data Availability

All data produced in the present work are contained in the manuscript

## Acknowledgements

GT is supported by the National Institute for Health Research (NIHR) Applied Research Collaboration South London at King’s College London NHS Foundation Trust, and by the NIHR Asset Global Health Unit award. The views expressed are those of the author(s) and not necessarily those of the NHS, the NIHR or the Department of Health and Social Care. GT is also supported by the Guy’s and St Thomas’ Charity for the On Trac project (EFT151101), and by the UK Medical Research Council (UKRI) in relation to the Emilia (MR/S001255/1) and Indigo Partnership (MR/R023697/1) awards

